# DETECTION AND SEGMENTATION OF ENDOSCOPIC ARTEFACTS AND DISEASES USING DEEP ARCHITECTURES

**DOI:** 10.1101/2020.04.17.20070201

**Authors:** Nhan T. Nguyen, Dat Q. Tran, Dung B. Nguyen

**Affiliations:** Medical Imaging Department, Vingroup Big Data Institute (VinBDI), Hanoi, Vietnam

## Abstract

We describe in this paper our deep learning-based approach for the EndoCV2020 challenge, which aims to detect and segment either artefacts or diseases in endoscopic images. For the detection task, we propose to train and optimize EfficientDet—a state-of-the-art detector—with different EfficientNet backbones using Focal loss. By ensembling multiple detectors, we obtain a mean average precision (mAP) of 0.2524 on EDD2020 and 0.2202 on EAD2020. For the segmentation task, two different architectures are proposed: UNet with EfficientNet-B3 encoder and Feature Pyramid Network (FPN) with dilated ResNet-50 encoder. Each of them is trained with an auxiliary classification branch. Our model ensemble reports an sscore of 0.5972 on EAD2020 and 0.701 on EDD2020, which were among the top submitters of both challenges.

## 1. INTRODUCTION

Disease detection and segmentation in endoscopic imaging play an important role in the early detection of numerous cancers, such as gastric, colorectal, and bladder cancers [1]. Meanwhile, the detection and segmentation of endoscopic artefacts is necessary for image reconstruction and quality assertion [2]. Many approaches [3, 4, 5] have been proposed to detect and segment artefacts and diseases in endoscopy. This paper describes our solution for the EndoCV2020 challenge, which consists of two tracks: one deals with artefacts (EAD2020) and the other one is for diseases (EDD2020). Each track is divided into two tasks: detection and segmentation. We tackle both tasks in both tracks by exploiting state-of-the-art deep architectures like EfficientDet [6] and U-Net [7] with variants of EfficientNet [8] and ResNet [9] as backbones. In the next sections, we provide a short description of the datasets, the details of the proposed approach, and experimental results.

## 2. DATASETS

EDD2020 [1] is a comprehensive dataset established to benchmark algorithms for *disease* detection and segmentation in endoscopy. It is annotated for 5 different disease classes, including BE, Suspicious, HGD, Cancer, and Polyp. The dataset comes with bounding boxes for disease detection and with masked image annotations for semantic segmentation. The training set includes total 386 endoscopy frames, each of which is annotated with either single or multiple diseases. Regions of the same class are merged into a single mask, while a bounding box of multiple classes is treated as separate boxes with the same location. Figure 1 shows the number of bounding boxes for each disease class.

**Fig. 1.**
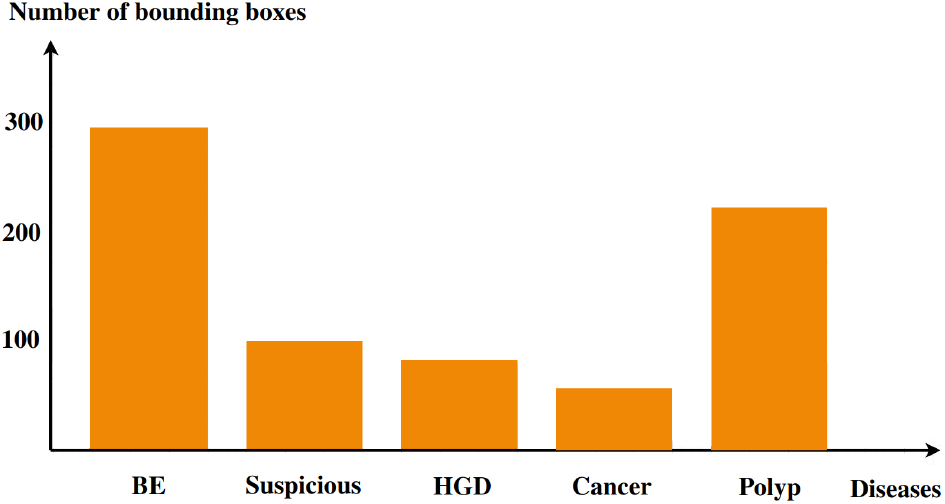
The number of bounding boxes for each disease class in training set provided by the EDD2020 dataset.

EAD2020 [10], on the other hand, is used for the track of endoscopy *artefact* detection and segmentation. The training set contains 2,531 annotated frames for 8 artefact classes, including specularity, bubbles, saturation, contrast, blood, instrument, blur, and imaging artefacts. Note that only first 5 classes are used for the segmentation task.

## 3. PROPOSED METHODS

### 3.1. Multi-class detection task

#### Detection network

For the detection task, we deployed EfficientDet [6], currently a state-of-the-art architecture for object detection. It employs EfficientNet [8] as the backbone network, BiFPN as the feature network, and shared class/box prediction network. Both BiFPN layers and class/box net layers are repeated multiple times based on different resource constraints. Figure 2 illustrates the EfficientDet architecture.

**Fig. 2.**
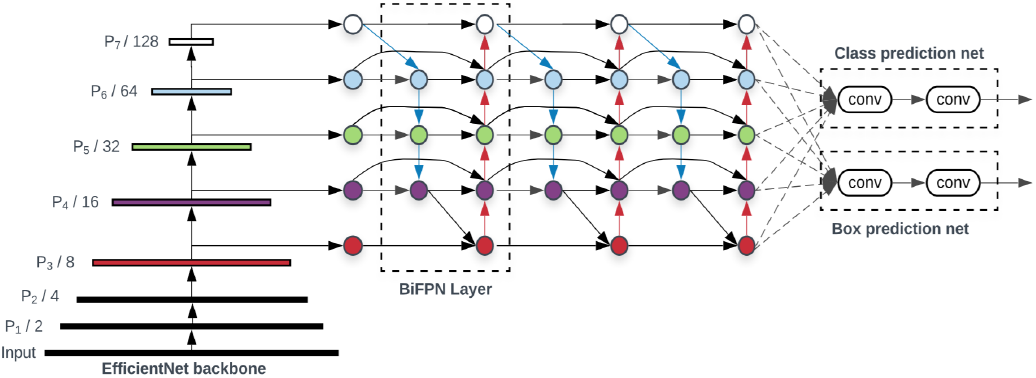
The EfficientDet architecture. The class prediction network was modified for providing the probabilities of 5 disease classes. The figure was reproduced from Tan *et al*. [6].

#### Training procedure

Due to the limited training data available (386 images in EDD2020 and 2531 images in EAD2020), we use various data augmentation techniques, including *random shift, random crop, rotation, scale, horizontal flip, vertical flip, blur, Gauss noise, sharpen, emboss*, and *contrast*. In particular, we found that the use of *mixup* could significantly reduce the overfitting. Given *x*_1_ and *x*_2_ as input images, the *mixup* image 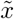 is constructed as

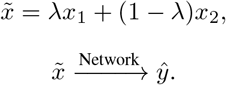

During training, our goal is to minimize the MixLoss *ℒ*_mixup_, which is expressed as

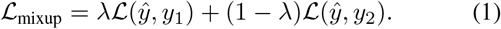

where the symbol *ℒ* denotes the Focal loss [11] and *λ* is drawn from *β*(0.75, 0.75) distribution; *y*_1_ and *y*_2_ are the ground-truth labels, while *ŷ* is the predicted label produced by the network. Fig. 3 visualizes a mixup example with *λ* being fixed to 0.5.

**Fig. 3.**
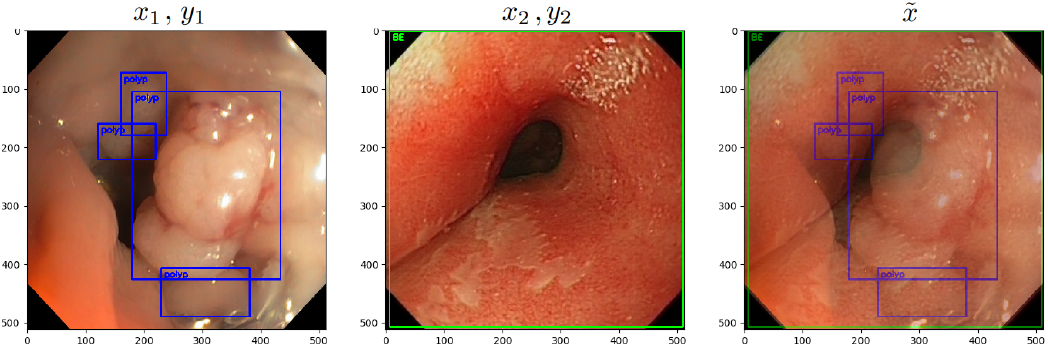
Mixup visualization with *λ* = 0.5.

Our detectors are optimized by the gradient decent using Adam update rule [12] with weight decay. In addition, cyclical learning rate [13] with restarts is also used. The ensemble of 6 models with different backbones (D0, D1, D2, D3, D4, and D5) using weighted box fusion [14] serves as our final model. Additionally, we search for the non-maximum suppression (NMS) threshold and the confidence threshold for different categories so that the resulting score (0.5 × mAP + 0.5 × IOU) is maximized.

### 3.2. Multi-class segmentation task

#### Segmentation network

We propose two different architectures for this task: U-Net with EfficientNet encoders and BiFPN with ResNet encoders.

#### U-Net

Our first network design makes use of U-Net with EfficientNetB3/B4 as backbones. We keep the original strides between blocks in EfficientNet and extract the feature maps from the last 5 blocks for the segmentation. A classification branch is used to provide the label predictions. The overall framework is depicted in Figure 4.

**Fig. 4.**
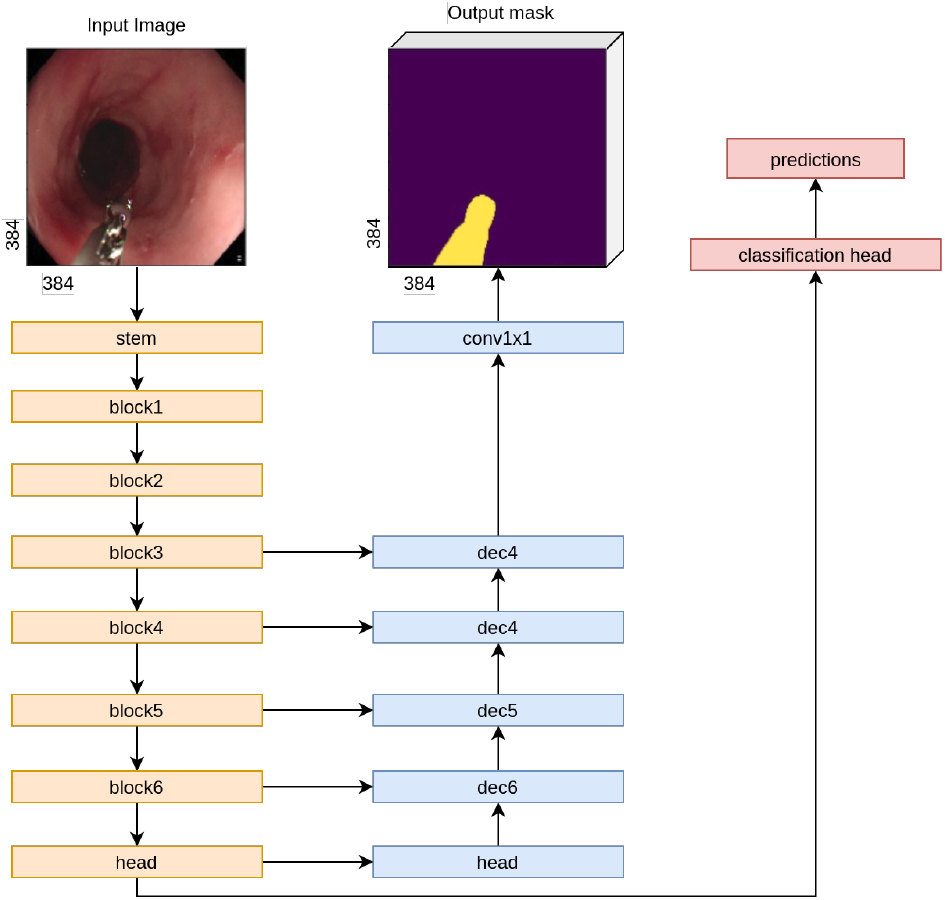
The U-Net with EfficientNetB3/B4 encoder and a classification branch architecture.

#### BiFPN

To generate the segmentation output from the BiFPN features, we combine all levels of the BiFPN pyramid by following the design illustrated in Figure 5. Starting with the deepest BiFPN level (stride-32 output), we apply three upsampling stages to obtain the feature map of the stride-4 output. An upsampling stage consists of a 3×3 Convolution, BatchNorm, ReLU and a 2× 2 bilinear upsampling. This strategy is repeated for other BiFPN levels with strides of 16, 8, and 4. The result is a set of feature maps at the same scale, which are then channel-wise concatenated. Finally, a 1× 1 Convolution, 4 ×4 bilinear upsampling and Sigmoid activation are used to generate the mask at the image resolution.

**Fig. 5.**
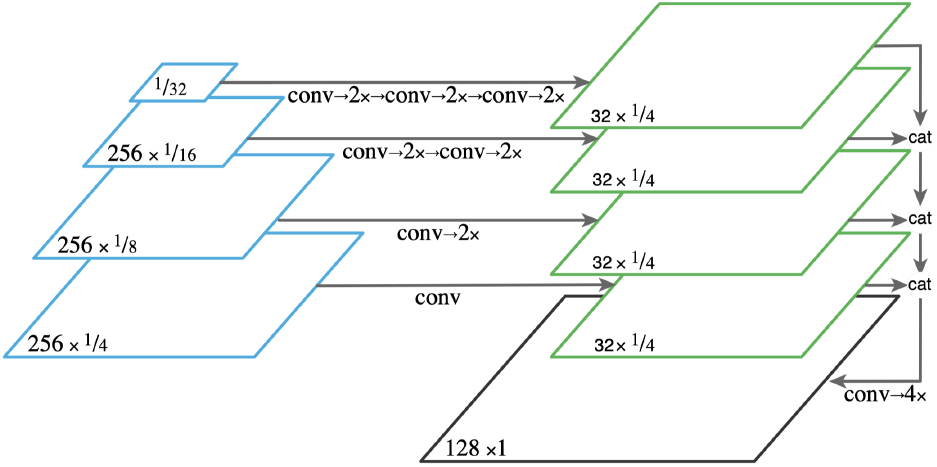
The BiFPN decoder for semantic segmentation.

#### Training procedure

All models are trained end-to-end with additional supervision from the multi-label classification task. The image labels are obtained directly from the segmentation masks. For example, if an image has B.E. mask annotation then the B.E. label is 1. Due to class imbalance in the training dataset, we use Focal loss for the classification task. Our final loss is *ℒ* = *ℒ*_seg_ +*λ*×*ℒ*_cls_ where *λ* = 0.4.

#### Inference

Relying solely on segmentation branch to predict masks will result in high false positives. Hence, we make use of the class predictions to remove masks. We search optimal classification thresholds to maximize the macro F1 score on the validation set. For every image, if the class probability is less than the optimal threshold then its predicted mask is completely removed.

## 4. EXPERIMENTAL RESULTS

Table 1 summarizes the detection and segmentation results of our submissions for both challenges. We describe the results of each sub-task below.

**Table 1.**
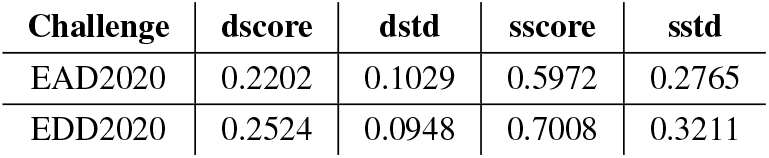
Detection and segmentation scores on the EndoCV2020 test set.

| Challenge | dscore | dstd | sscore | sstd |
| --- | --- | --- | --- | --- |
| EAD2020 | 0.2202 | 0.1029 | 0.5972 | 0.2765 |
| EDD2020 | 0.2524 | 0.0948 | 0.7008 | 0.3211 |

Results on the validation set of EDD2020 for the detection task are detailed in Table 2. Our best single model (*i*.*e*. EfficientDet-D5) obtained a detection score (dScore) of 0.41. The best detection performance was provided by the ensemble model, which reported a dScore of 0.44, a mean mAP of 0.36 ± 0.05, and an IoU of 0.52. As shown in Table 1, our ensemble model yielded dScores of 0.2524 ± 0.0948 and 0.2202 ± 0.1029 on the hidden test sets of EDD2020 and EAD2020, respectively.

**Table 2.** Experimental results on EDD2020 validation set.

| Method | dScore | mAP | IoU |
| --- | --- | --- | --- |
| ED0 [6] | 0.23 | $0.13 \pm 0.04$ | 0.33 |
| ED0, Augs | 0.34 | $0.26 \pm 0.07$ | 0.42 |
| ED0, Augs, Mixup, CLR [15] | 0.40 | $0.30 \pm 0.05$ | 0.51 |
| ED5, Augs, Mixup, CLR [15] | 0.41 | $0.29 \pm 0.05$ | 0.54 |
| Ensemble (ED0-ED5), WBF [14] | 0.44 | $0.36 \pm 0.05$ | 0.52 |

Results on validation sets for the segmentation task are provided in Table 3 and Table 4. On the EDD2020 validation set, our best single model achieved a Dice score of 0.854 and an IoU of 0.832. On the EAD2020 validation set, we obtained a Dice score of 0.732 and an IoU of 0.578. As shown in Table 1, our ensemble achieved a segmentation score (sscore) of 0.5972 in the EAD2020 challenge and an sscore of 0.7008 in the EDD2020 challenge, both of which were among the top results for the segmentation task of both tracks.

**Table 3.** 5-fold cross-validation results on EDD2020.

| Method | Dice | IoU |
| --- | --- | --- |
| UNet-EfficientNetB4 [8][7] | $0.8522 \pm 0.0221$ | $0.8279 \pm 0.0213$ |
| BiFPN-ResNet50 | $0.8544 \pm 0.0232$ | $0.8317 \pm 0.0228$ |

**Table 4.** 3-fold cross-validation results on EAD2020.

| Method | Dice | IoU |
| --- | --- | --- |
| UNet-EfficientNetB4 | $0.7131 \pm 0.0379$ | $0.555 \pm 0.0451$ |
| BiFPN-ResNet50 | $0.7325 \pm 0.0162$ | $0.578 \pm 0.0201$ |

## 5. CONCLUSION

We have described our solutions for the detection and segmentation tasks on both tracks of EndoCV2020: EAD for artefacts and EDD for diseases. By using EfficientDet for detection and U-Net/BiFPN for segmentation, we obtained significant results on both datasets, especially for the segmentation task. These results suggest that some of the deep architectures that are effective for natural images can also be useful for medical images like endoscopic ones, even with a small-size training datasets.

## Data Availability

The data that support the findings of this study are openly available at EDD2020 webpage.

https://ieee-dataport.org/competitions/endoscopy-disease-detection-and-segmentation-edd2020

## Notes

### Competing Interest Statement

The authors have declared no competing interest.

### Funding Statement

This work was supported by the Vingroup Big Data Institute (VinBDI), Hanoi, Vietnam.

